# Improving Machine Learning Prediction of ADHD Using Gene Set Polygenic Risk Scores and Risk Scores from Genetically Correlated Phenotypes

**DOI:** 10.1101/2022.01.11.22269027

**Authors:** Eric Barnett, Yanli Zhang-James, Stephen V Faraone

## Abstract

**Background:** Polygenic risk scores (PRSs), which sum the effects of SNPs throughout the genome to measure risk afforded by common genetic variants, have improved our ability to estimate disorder risk for Attention-Deficit/Hyperactivity Disorder (ADHD) but the accuracy of risk prediction is rarely investigated.

**Methods:** With the goal of improving risk prediction, we performed gene set analysis of GWAS data to select gene sets associated with ADHD within a training subset. For each selected gene set, we generated gene set polygenic risk scores (gsPRSs), which sum the effects of SNPs for each selected gene set. We created gsPRS for ADHD and for phenotypes having a high genetic correlation with ADHD. These gsPRS were added to the standard PRS as input to machine learning models predicting ADHD. We used feature importance scores to select gsPRS for a final model and to generate a ranking of the most consistently predictive gsPRS.

**Results:** For a test subset that had not been used for training or validation, a random forest (RF) model using PRSs from ADHD and genetically correlated phenotypes and an optimized group of 20 gsPRS had an area under the receiving operating characteristic curve (AUC) of 0.72 (95% CI: 0.70 – 0.74). This AUC was a statistically significant improvement over logistic regression models and RF models using only PRS from ADHD and genetically correlated phenotypes.

**Conclusions:** Summing risk at the gene set level and incorporating genetic risk from disorders with high genetic correlations with ADHD improved the accuracy of predicting ADHD. Learning curves suggest that additional improvements would be expected with larger study sizes. Our study suggests that better accounting of genetic risk and the genetic context of allelic differences results in more predictive models.

## Introduction

The field of psychiatric genomics has made great strides discovering genetic loci that are significantly associated with psychiatric disorders [1-3]. These discoveries have generated new hypotheses about the genomic architecture complex pathogenesis of many of these disorders. The combination of risk conferring alleles has improved the prediction of psychopathology [4].

A multi-site ADHD GWAS found that 12 genome-wide-significant loci captured a small amount of the heritability of ADHD while risk profiles using all loci captured a significantly larger amount of heritability, which proved the usefulness of loci that are, individually, are not significantly different between cases and controls [1]. Even in this study of over 20,000 people with ADHD, the complex genetic architecture of the disorder makes predicting generalizable risk and establishing significance at each common variant difficult.

Previous work has shown that ADHD has significant genetic overlaps with other psychiatric and non-psychiatric disorders [5-10]. This supports the theory that ADHD risk comprises traits that are also present in the phenotypes with which it is genetically correlated. The risk estimation of SNPs in genetically correlated disorders could be more predictive in ADHD relative to the risk estimation of SNPs in ADHD GWASs due to larger sample sizes being better for estimating risk. In addition, when dealing with disorders with high heterogeneity like ADHD it is possible that other less heterogeneous phenotypes better estimate risk for genetic loci for some clusters of patients. Therefore, using the genetic overlap with other disorders could be useful in improving the predictive modeling of ADHD.

A review of twin-studies of ADHD found that the mean heritability of ADHD across 37 studies was 74% [11]. The high heritability of ADHD suggests that predicting ADHD using genetic and environmental data is achievable. However, reports on predictive models of ADHD using genetic data is limited. Significant improvements in prediction and our understanding of the disorder must be made before genetic information can be used in the clinic as part of future objective diagnoses and personalized medicine plans that aim to improve outcomes in ADHD.

One potential area of improvement is balancing the flexibility of models to detect robust risk patterns with complexity and generalizability. Combining the risk at SNPs across the genome into a single polygenic risk score (PRS) has proven to be a successful way to create a more useful and generalizable feature than any individual SNP[12]. However, summing all SNPs into a single value per individual limits any modelling method’s capacity to learn more complicated patterns and interactions. On the other end of the complexity spectrum, using individual SNPs as input into machine learning models of complex and heterogenous disorders like ADHD leads to concerns of overfitting and lack of generalizability [13]. Combining risk at the gene set level could be an effective middle ground between these two extremes. While research using features combining risk at the gene set level to predict a disorder is limited, gene set association analyses have shown that this middle ground can be useful.

While machine learning classification models of ADHD using genomic data have not been reported, many researchers have used such models to predict diagnoses for other heritable complex disorders[14-18]. Collectively, these studies have shown the potential of machine learning to predict many disorders but concerns of how well these models would perform on unseen external data sets remain. In addition, many machine learning methods generate “black-box” models that are uninterpretable. Since most models lack the performance necessary for clinical application, interpretable models may provide additional useful results apart from the model that would otherwise only be an intermediate to eventual models that will be useful clinically. Interpretable genomic models could yield biological insights by finding new loci of interest or new groups of loci that together improve models. These models also could incorporate further model validation by relating the output to our understanding of the biology behind the disorder.

Here, we balance these issues by summing risk across gene sets to create gene set polygenic risk scores (gsPRSs) that may be used alongside PRS to improve predictive accuracy by providing the model with information about gene sets associated with ADHD. We hypothesized that including gsPRSs as input into machine learning models would improve prediction performance compared to models that use only traditional PRS. We also supplemented the model with summary statistics from phenotypes with high genetic correlations with ADHD as additional features to test if these information are useful to improve ADHD prediction.

## Methods

### Data Preprocessing and Splitting

Quality control and imputation were done using the RICOPILI pipeline[19]. After quality control, 2455 ADHD cases and 8432 controls across 9 cohorts aggregated by the PGC were available for analysis [1]. We excluded SNPs with a minor allele frequency < 0.01, missing genotype rate

> 0.05, and deviating from Hardy-Weinberg equilibrium in controls at p < 1 × 10^−5^. The participants were randomly split into a training subset containing 1673 cases and 5818 controls, a validation subset containing 406 cases and 1329 controls, and a test subset containing 376 cases and 1285 controls. The training subset was used to teach the model to differentiate different cases and controls by optimizing the parameters within the model. The validation subset was used to estimate the model performance outside examples used to train the model and to optimize model hyper-parameters. The test subset was used for reporting the results of our final models on an unseen sample.

### Gene Set Association Analysis

Using the SNP association p-values generated in the SNP association analysis, we used MAGMA to compare allele frequencies between cases and controls at the gene and gene sets level [20]. Both analyses used an extended gene window starting 35 kilobases upstream and ending 10 kilobases downstream of each gene to account for cis regulatory elements. The complete MsigDB gene ontology gene sets collection was used as input into the analysis. The gene sets most associated with this study sample have been previously reported [1].

### Polygenic Risk Scoring

From the associations collected from gene set analysis, we selected the most associated gene sets based on their p-values. To avoid including the same risk signal multiple times within a score, we adjusted SNPs tagging each gene set for linkage disequilibrium using PRS-CS, a tool that infers posterior effect sizes of each SNP after removing overlaps due to linkage disequilibrium. From these adjusted SNPs, we used polygenic weighted scoring to generate a risk profile for each gene set in each subject using Plink. We calculated genome-wide polygenic risk profiles using the same combination of PRS-CS and Plink scoring. For comparison, we also generated PRS using the clumping and thresholding method.

### Correlated Trait/Disorder Polygenic Risk Scoring

We calculated additional risk profiles using SNP effects estimated from GWASs of disorders and traits with the highest genetic correlation with ADHD and heritability over 0.1 found using GWAS Atlas[21]. After excluding similar phenotypes based on study size, the included phenotypes were age at first sexual intercourse[21], opioid use[22], college completion[23], childhood IQ[24], childhood extreme obesity[25], autism spectrum disorder[26], time spent watching television[21], psychiatric cross-disorder risk[27], intracranial volume[28], age at menopause[21], and myopia[21]. We calculated the gsPRS for genetically correlated disorders on the gene sets most associated with ADHD diagnosis by using the SNP effects from the summary statistics for each trait in additional MAGMA gene set analyses. We included PRS and 100 gsPRS for each trait/disorder in machine learning feature selection.

### Machine Learning Preprocessing and Feature Selection

We adjusted each polygenic risk score for ancestry by extracting the top 5 principal components from a principal components analysis (PCA) of the training subset and using those 5 principal components in a generalized linear model predicting each polygenic risk score. We replaced the unadjusted polygenic risk score with the residual of each prediction using the 5 principal components. We normalized each score between 0 and 1 using min-max normalization and balanced cases and controls in each subset by random case up-sampling with replacement.

For gsPRS only models, we started by selecting gsPRS from the 40 gene sets most associated with ADHD within the training subset. We optimized the hyperparameters of a random forest based on this initial set of features. Then, we performed a random iterative feature selection process in which we kept and recorded the most important features, based on the permutation feature importance calculated from the mean difference in Gini impurity, and randomly replaced the less important features with a different gsPRS feature until the model found a set of gsPRS that outperformed the previous best set. We reoptimized the random forest hyperparameters at regular intervals and repeated the random replacement process such that each feature would likely be included in multiple iterations of the newly optimized model. At the end of this process, we selected the best group of 20 gsPRS for model performance evaluation.

In the models that included gsPRS and PRS-CS, we used the same random iterative feature selection approach used in the gsPRS only model, but also included the genome-wide PRS-CS scores calculated from the training subset and summary statistics from GWAS of related disorders in every model.

### Machine Learning Model Optimization

Within Scikit-learn, we used grid search optimization to select the best hyperparameters for all models using the AUC in the validation subset [29]. We optimized multiple types of models to better compare the performance of different methods within the validation subset and select the best model for this application. Exploring multiple models is essential given that for any given problem, one algorithm may be ideal but it is not possible, in advance, to know what algorithm will be best [30]. For random forest (RF) models, we optimized number of trees in the forest, maximum depth of the tree, and the number of features to consider when looking for the best split. For support vector machine (SVM) models, we optimized C, which balances misclassification against simplicity, and gamma, which determines the effect of a single training example on the model. For k-nearest neighbor (kNN) models, we optimized number of neighbors, leaf size, weight function, and the power parameter for the Minkowski metric. For the PRS and PRS-CS models, we fit logistic regression models to compare performance with our more complex models using the glm package in R. We fit the lasso model using all PRS-CS and all gsPRS as input using the glmnet package in R.

### Model Performance Evaluation and Feature Importance Tracking

To measure the performance of the models selected with grid search optimization we used area under the receiver operating characteristic curve (AUC) in the test subset. Data leakage is a common issue in machine learning research normally caused by inadvertently learning information about the test data that improves performance in those specific data. One way data leakage can occur is through testing many models on the test data, which increases the chance of selecting a model that is randomly configured in a way that is more optimal for the test data but not generalizable. With the goal of minimizing data leakage that might bias our results towards the test data, we tested model performance in the test subset only on the model with the highest AUC in the validation subset for each analysis. We estimated the known genetic variance explained by each of the models using a formula developed for the genetic interpretation of AUCs using 0.75 as the heritability estimate and 0.05 as the prevalence estimate[31]. We compared AUCs from different models using DeLong’s test for two correlated ROC curves. We also tested the probability of achieving the AUC in the best gsPRS grouping by comparing the AUC in the test subset with the distribution of AUCs from 10,000 models with random gsPRS groups of the same size. All models included the PRS for all correlated phenotype summary statistics. We used learning curves to model whether additional training examples would improve model performance and to compare models.

To calculate a more generalizable importance score for each gsPRS outside of the best group of gsPRS, we estimated feature importance for each gsPRS and PRS-CS feature in RF models with a random group of gsPRS calculated from gene sets associated with ADHD and tracked the permutation feature importance that measures the decrease in model performance when a single feature value is randomly shuffled. We calculated the mean feature importance of each gsPRS across 10,000 models that used 40 random gsPRSs each. We did not use feature importance scores calculated from the test subset for feature selection or any optimization.

### Testing Biological Relevance of gsPRS Feature Importance

To further validate our methods by testing for correlations with the known neurobiology of ADHD, we computed correlations between tissue-specific gene expression and feature importance [32, 33]. For ADHD, we would expect that most gene sets truly associated with the disorder would be more relevant to the brain and less relevant to other tissues. Therefore, if the importance of the gsPRS generated in our analysis are correlated with brain expression relative to all other tissues, we can be more confident that gsPRSs are collectively picking up a real generalizable risk feature instead of modelling random noise. We used a dataset containing gene expression data for 54 tissue types from the genotype-tissue expression (GTEx) project. We combined this gene expression data into gene set expression data for the same gene sets used in the gsPRSs. We estimated relative gene set expression in the brain using the Preferential Expression Measure formula which estimates how different the expression of a gene is relative to the expected expression level. We fit a linear model predicting gene set expression in brain tissues relative to non-brain tissues using the MAGMA gene set association p-value to establish a baseline. We fit a linear model using the base model with gsPRS feature importance as a second predictor to test for association of gene set expression with gsPRS importance score after controlling for MAGMA gene set associations. We fit linear models with the same dependent and independent variables using only gene sets calculated from the ADHD training subset or from the group of correlated phenotypes to test whether each group was independently associated with gene set expression. To test whether the association between mean importance score and relative gene set brain expression in the brain was dependent on whether the gsPRS was calculated from the ADHD training subset, we estimated predictive margins using STATA16’s margins command, which computes the average probability for each observation at a fixed level of a selected variable. In our analyses, these predictive margins estimate the average relative gene set expression in the brain for each gsPRS while fixing the ADHD vs non-ADHD variable to each value. A meta-analysis on subcortical brain volume differences in ADHD found that the volumes of the accumbens, amygdala, caudate, hippocampus, and putamen were smaller in participants with ADHD [34]. We fit linear models predicting gene expression in these brain regions implicated in ADHD relative to all other brain regions with gene set expression as the dependent variable and MAGMA gene set association p-value and gsPRS feature importance.

## Results

### Model performance

To establish baseline performance, we measured the prediction performance in the test subset of a logistic regression with the PRS calculated from the training subset. This PRS only logistic regression had an AUC of 0.62 (95% CI: 0.60 – 0.64) in the test subset and explained 5.0% of the known genetic variance. Replacing PRS with PRS-CS in another logistic regression model led to an AUC of 0.66 (Figure 1; 95% CI: 0.64 – 0.68) and explained 9.0% of the known genetic variance. We then measured the performance of logistic regression and random forest models containing the PRS-CS from the training subset and PRS-CS calculated from summary statistics from phenotypes with a heritability above 0.1 with the highest genetic correlation to ADHD (Table 1). The logistic regression model had an AUC of 0.66 (95% CI: 0.64 – 0.68) while a random forest model using the same input had an AUC of 0.69 (Figure 1; 95% CI: 0.67 – 0.71) in the test subset and explained 12.8% of the known genetic variance.

**Figure 1.**
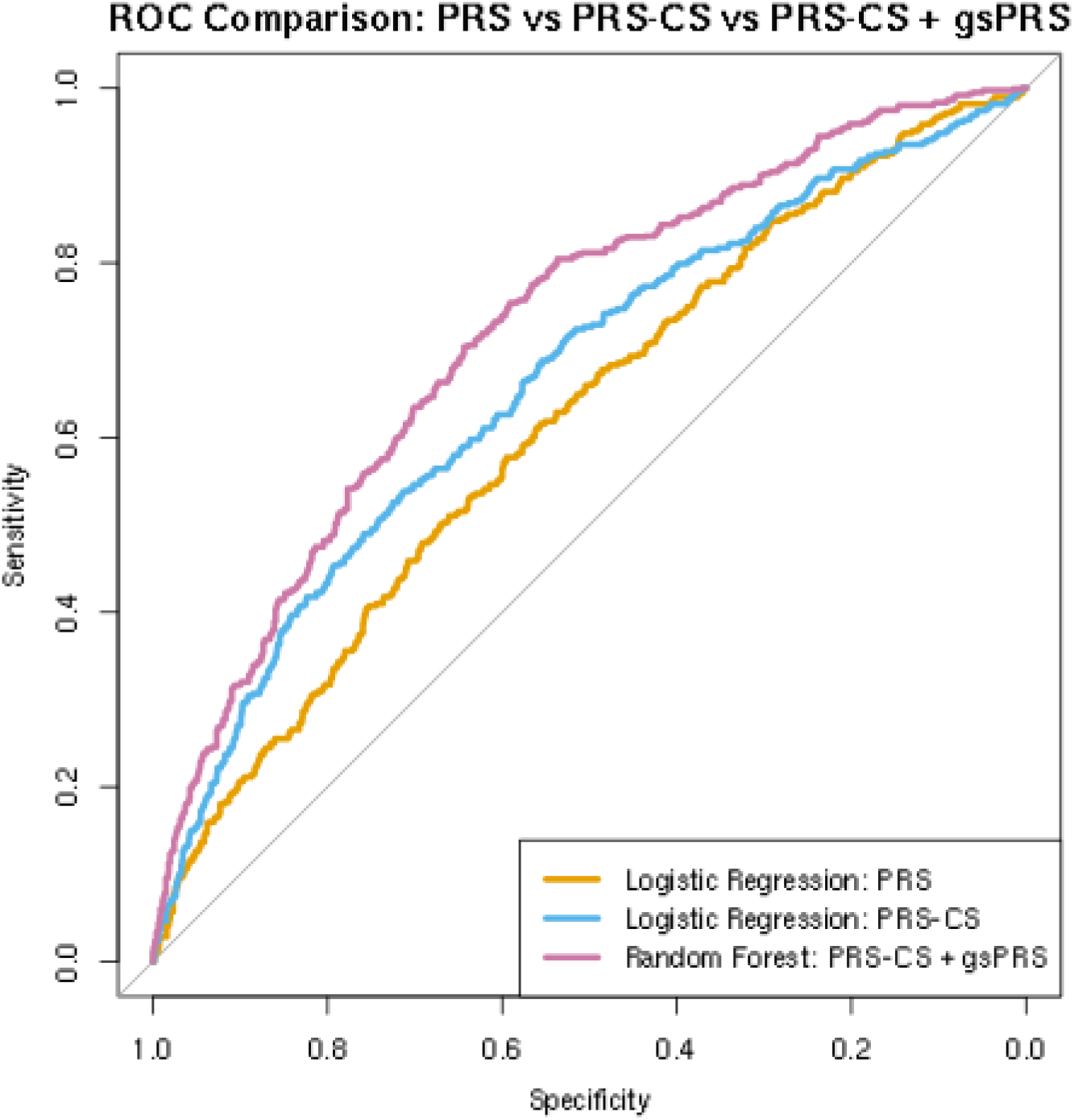
Model ROC Comparison. The logistic regression model using traditional PRS methods had an AUC of 0.62 (95% CI: 0.60 – 0.64). The logistic regression model using PRS-CS methods had an AUC of 0.66 (95% CI: 0.64 – 0.68). The random forest model using PRS-CS and an optimized group of 20 gsPRS had an AUC of 0.72 (95% CI: 0.70 – 0.74).

**Table 1.** Summary statistics from genetically correlated phenotypes that were used to generate additional genetic risk features.

| Trait |  | rg | N | SNP.h2 |
| --- | --- | --- | --- | --- |
| Age at first sexual intercourse |  | -0.584 | 339614 | 0.1132 |
| Opioid use |  | 0.565 | 78808 | 0.146 |
| College completion |  | -0.524 | 95427 | 0.105 |
| Childhood IQ |  | -0.461 | 12441 | 0.2744 |
| Extreme obesity (childhood) |  | 0.436 | 7916 | 0.5078 |
| Autism spectrum disorder |  | 0.384 | 46350 | 0.1944 |
| Time spent watching television (TV) |  | 0.372 | 365236 | 0.1023 |
| PGC cross disorder |  | 0.262 | 61220 | 0.1715 |
| Intracranial Volume |  | -0.248 | 26577 | 0.2467 |
| Myopia |  | -0.217 | 78647 | 0.1532 |

After using our feature selection method to select the best group of 20 gsPRS, we trained a random forest model using the selected group and all PRS-CS. In the test subset, this model had an AUC of 0.72 (Figure 1; 95% CI: 0.70 – 0.74) and explained 17.4% of the known genetic variance. This was a significant improvement in comparison to the RF that included only the PRS-CS from each trait (p = 0.0057, DeLong’s test for two correlated ROC curves). The RF model with all PRS-CS and the best group of 20 gsPRS also had a significantly higher AUC (p = 1.2 × 10^−6^, Delong’s test for two correlated ROC curves). compared to a lasso model fit with all PRS-CS and gsPRS as input, which had an AUC of 0.65 (95% CI: 0.63 – 0.67). The AUC of the best group model was greater than 99.6% of the 10,000 random group models. The mean AUC of the random group models was 0.69. All the gene sets used in the random groups were associated with ADHD in the training subset with a p-value of less than 0.05 without correction for multiple testing. SVM and kNN models were less predictive than RF models in the validation subset, so we did not test them on the test subset.

We trained and optimized another random forest model using only gsPRS. The model had an AUC of 0.61 (95% CI: 0.59 – 0.63) in the test subset. The AUC of the best group model was greater than 99.1% of the 10,000 random group models.

### Random Forest Learning Curve and Feature Importance Analyses

For the best random forest model, we generated a learning curve (Figure 2) that plots the AUC against the number of training examples[35]. We also optimized a random forest model using only PRS-CS and generated a learning curve (Figure 3) for comparison.

**Figure 2.**
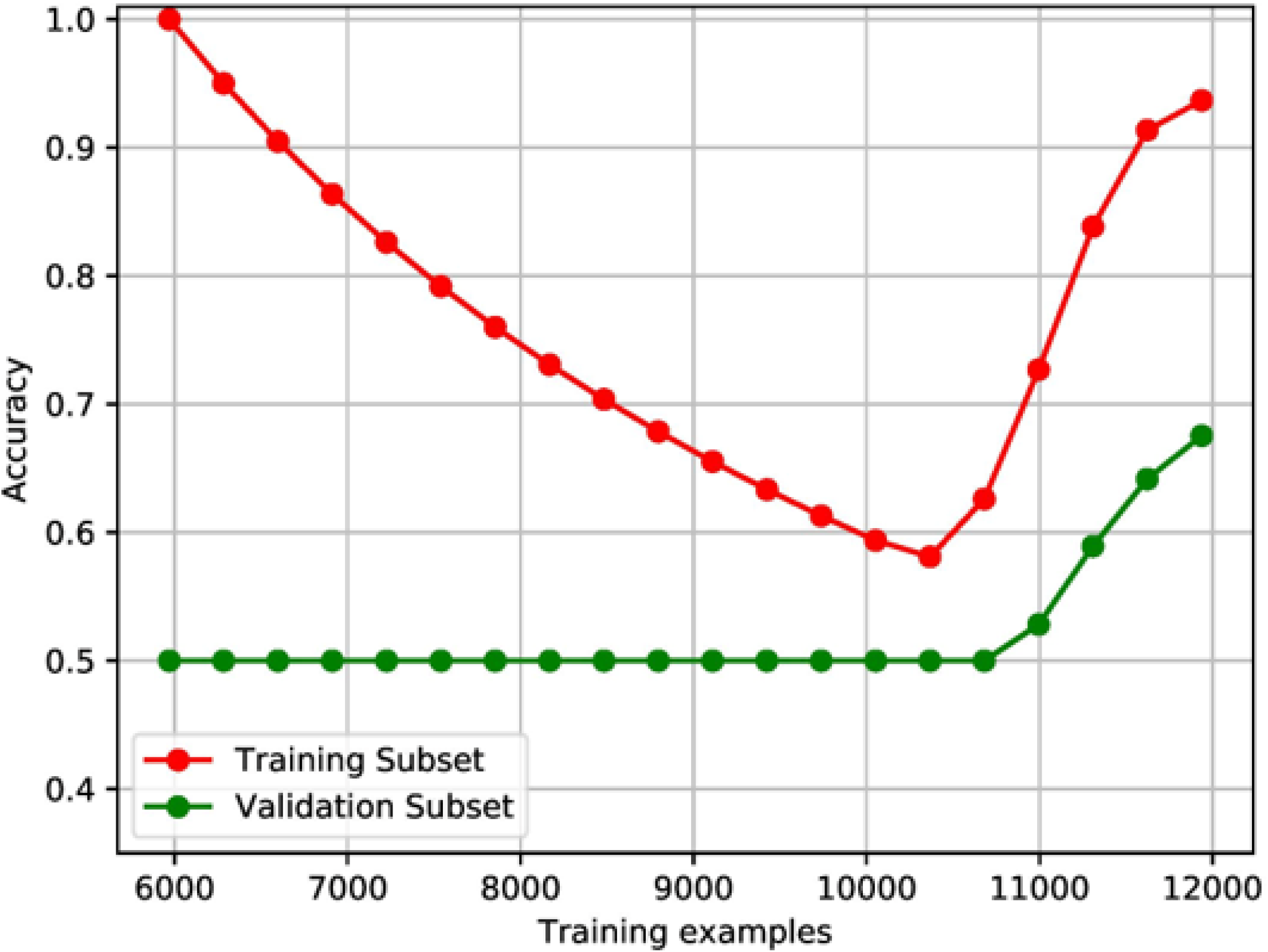
Learning Curves for gsPRS + PRS-CS Random Forest Model. The learning curve analysis of the random forest model containing all PRS-CS and the best group of 20 gsPRS. Each point represents the accuracy of the model when trained with a set number of training examples.

**Figure 3.**
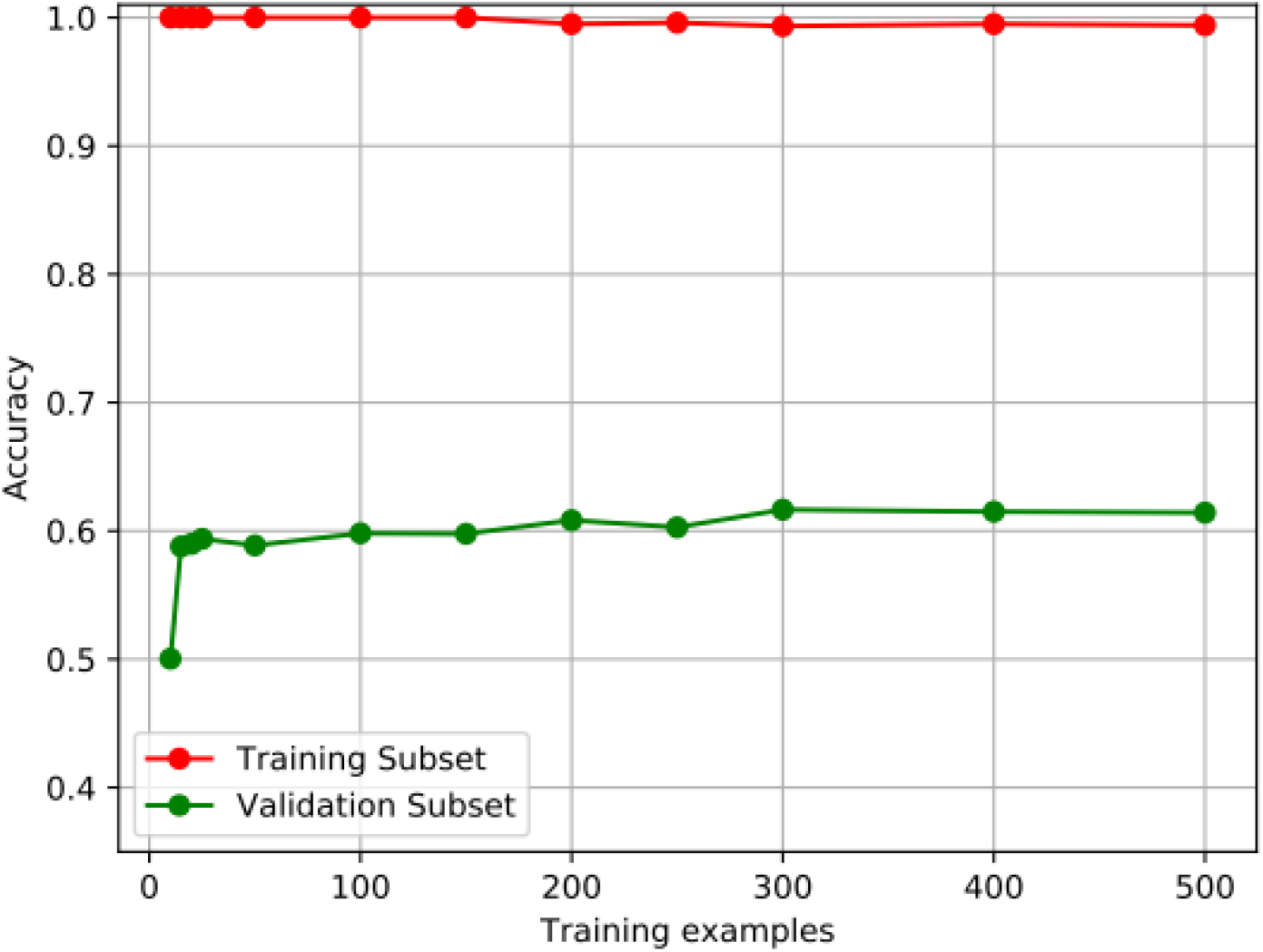
Learning Curves for PRS-CS only Random Forest Model. The learning curve analysis of the random forest model containing all PRS-CS. Each point represents the accuracy of the model when trained with a set number of training examples.

Using the optimized random forest model, we generated feature importance scores in the test subset for all the features used in the model. The most important features and their importance scores are listed in Table 2. In addition, we calculated the average feature importance in the test subset across 10,000 random group of 40 gsPRS only models (princTable 1).

**Table 2.** Top feature importance scores for the best group of gsPRS and PRS random forest model.

| Feature | Phenotype | Importance |
| --- | --- | --- |
| Genome Wide Polygenic Risk Score | pgc cross disorder | 0.0324 |
| Genome Wide Polygenic Risk Score | training subset | 0.0250 |
| GO_CELLULAR_RESPONSE_TO_ENDOGENOUS_STIMULUS | training subset | 0.0055 |
| GO_POSITIVE_REGULATION_OF_PROTEIN_MODIFICATION_PROCESS | training subset | 0.0044 |
| GO_RESPONSE_TO_TOXIC_SUBSTANCE | training subset | 0.0032 |
| GO_REGULATION_OF_PRESYNAPSE_ORGANIZATION | training subset | 0.0027 |
| GO_REGULATION_OF_PLASMA_LIPOPROTEIN_PARTICLE_LEVELS | training subset | 0.0026 |
| GO_POSITIVE_REGULATION_OF_TRANSFERASE_ACTIVITY | training subset | 0.0026 |
| GO_PRESYNAPSE_ORGANIZATION | training subset | 0.0025 |
| GO_SYNAPTIC_SIGNALING | pgc cross disorder | 0.0024 |
| Genome Wide Polygenic Risk Score | age first had sexual intercourse | 0.0022 |
| GO_REGULATION_OF_VASCULAR_PERMEABILITY | training subset | 0.0019 |
| GO_PRIMARY_ALCOHOL_BIOSYNTHETIC_PROCESS | training subset | 0.0018 |
| GO_HEAD_DEVELOPMENT | pgc cross disorder | 0.0017 |
| GO_LIPASE_ACTIVITY | training subset | 0.0017 |
| GO_REGULATION_OF_NEURON_DIFFERENTIATION | pgc cross disorder | 0.0014 |
| GO_MICROTUBULE_PLUS_END_BINDING | training subset | 0.0013 |
| GO_NEURON_DIFFERENTIATION | pgc cross disorder | 0.0012 |
| GO_CEREBELLAR_GRANULAR_LAYER_FORMATION | training subset | 0.0011 |
| GO_SOMATODENDRITIC_COMPARTMENT | pgc cross disorder | 0.0009 |
| GO_POSITIVE_REGULATION_OF_EXCITATORY_POSTSYNAPTIC_POTENTIAL | training subset | 0.0008 |
| GO_REGULATION_OF_MEMBRANE_POTENTIAL | pgc cross disorder | 0.0008 |

### Testing Biological Relevance of gsPRS Feature Importance

The base linear model we fit with relative gene set expression as the dependent variable and MAGMA gene set association p-value as the independent variable showed a significant negative correlation between the two variables (p = 1 × 10^−5^). The model adding mean gsPRS importance score as an independent variable showed a significant positive correlation between mean gsPRS importance score and relative gene set expression after controlling for MAGMA gene set association p-value (p = 2 × 10^−4^). We found no significant differences in gene expression between brain regions implicated in ADHD and other brain regions.

The base + mean gsPRS feature importance model we fit using only gsPRS calculated from the ADHD training subset showed a significant positive correlation between mean gsPRS importance score and relative gene set expression in the brain (p = 0.008). The same model fit using only gsPRS calculated from the correlated phenotypes also showed a significant positive correlation between mean gsPRS importance score and relative gene set expression in the brain (p = 0.003). An additional linear model we fit adding an independent variable specifying whether the gsPRS was calculated in the ADHD training subset or a correlated phenotype and that variable’s interaction with importance score showed that the correlation of gene set expression in the brain with mean gsPRS importance was negatively dependent on whether the gsPRS was calculated in the ADHD training subset (p = 5 × 10^−4^). As illustrated in Figure 4, our predictive margins analysis of this interaction estimated a significant positive association with a slope of 4.7 (p < 0.001) when the variable indicating development in the ADHD training subset was fixed to 0, meaning the gsPRS was developed using one of the correlated phenotypes, and a significant positive association with a slope of 0.60 (p = 0.015) when the same variable was fixed to 1, meaning the gsPRS was developed using the ADHD training subset.

**Figure 4.**
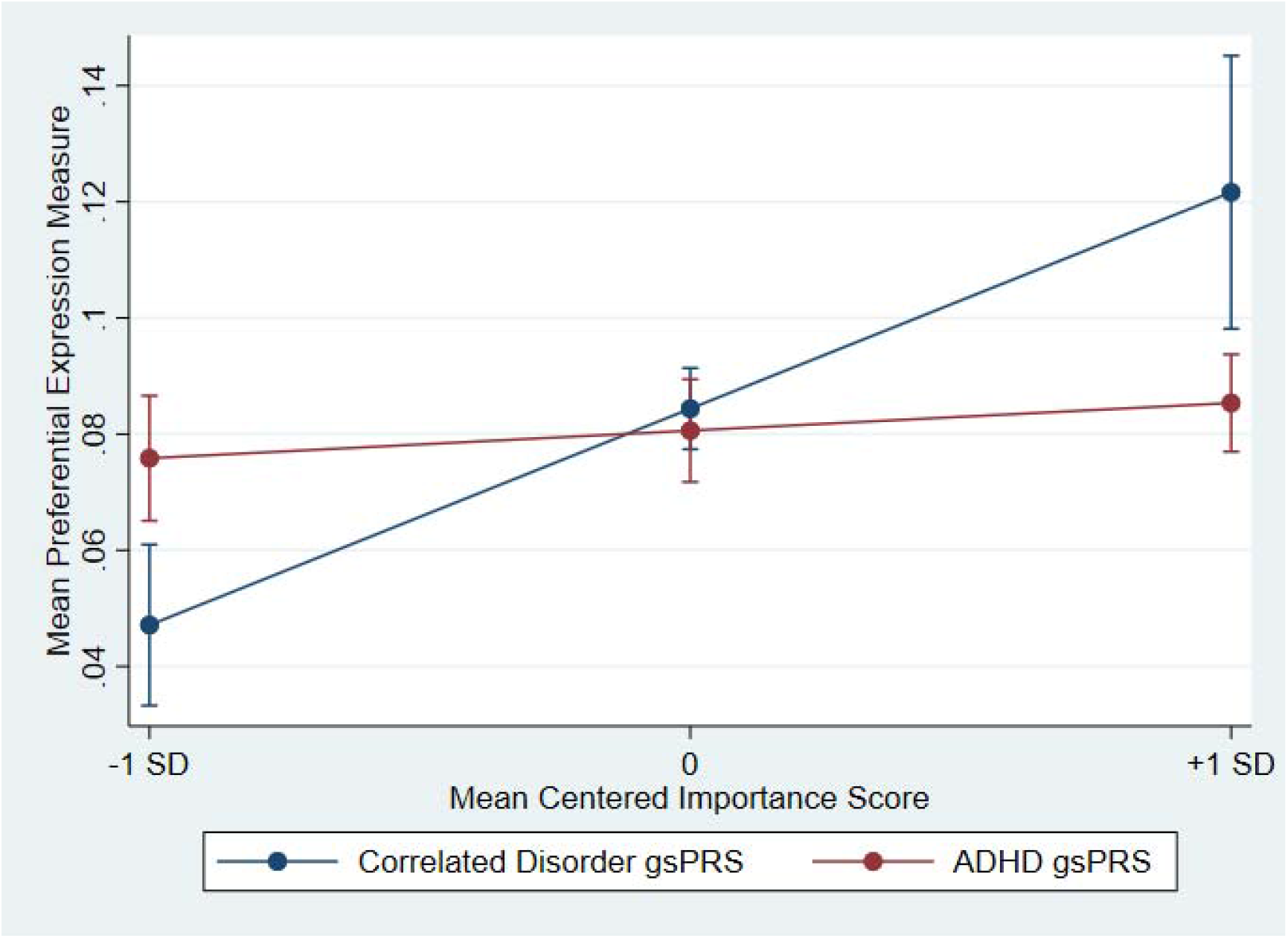
Predictive Margins Analysis of Interaction between Importance Score and Relative Gene Set Expression. When the variable indicating gsPRS development in the ADHD training subset was fixed to 0 (developed in a correlated disorder) there was a significant positive association with a slope of 4.7 (p < 0.001). When the variable was fixed to 1 (developed in the ADHD training subset) there was a significant positive association with a slope of 0.60 (p = 0.015).

## Discussion

This study is the first to produce gene set specific risk profiles predicting the presence/absence of a psychiatric disorder with machine learning. The addition of optimized groups of gsPRS to genome wide PRS-CS significantly improves prediction performance compared to both models without gsPRS and models with random groups of gsPRS. We further validated these results by testing for biological correlation of the random forest importance scores, which showed that importance scores were significantly positively associated with increased relative gene set expression in the brain.

Compared to simpler models that rely on a single PRS value per individual and more complex models that rely on “black-box” dimension reduction methods, gsPRS models have the potential of offering more interpretability and have the possibility to shed light on mechanisms involved in risk prediction and test specific gene set hypotheses. To improve interpretation of our models, we generated two sets of feature importance measurements that capture similar, but distinct information regarding the predictiveness of gsPRS. The feature importance measurements from the best group of gsPRS (Table 2) show how useful each gsPRS and PRS-CS were in that specific model. Unsurprisingly, the ranking is led by the PRS-CS from a cross-disorder GWAS that studied the shared risk across multiple psychiatric disorders including ADHD and the PRS-CS calculated from the training subset. Those PRS-CS are followed by a group of gsPRS that collectively led to significant improvements in prediction. It is likely that this group contains less overlapping risk information relative to other groupings since such overlaps would increase model complexity without adding value for prediction. However, overlapping gsPRS could still be important individually or in different groupings. Therefore, we calculated average gsPRS feature importance in 10,000 models that each used 40 gsPRS as input. This average represents how often and how strongly each gsPRS was able to improve prediction.

With this list of gsPRSs and their feature importance, we sought to further validate our methods by testing for correlations with what is known about the neurobiology of ADHD [32, 33]. Our baseline regression analysis found a significant negative correlation between relative gene set expression in the brain and MAGMA gene set association p-value. This met our expectation since MAGMA is a widely used tool and we would expect that gene sets more associated with ADHD and correlated phenotypes would be correlated with increased relative expression of that gene set in the brain, which is consistent with the report of Demontis et al [1]. Our analysis adding mean gsPRS importance score to the baseline regression analysis found that, even after correcting for MAGMA gene set association, mean gsPRS importance score was significantly positively correlated with relative gene set expression in the brain. This suggests that the mean gsPRS importance scores can be used to select biologically relevant gene sets beyond their association with ADHD as calculated using MAGMA. This finding suggests that combining MAGMA and mean gsPRS importance scores could provide a better way to prioritize gene sets for future study compared with using MAGMA alone.

We were also interested to test whether the correlations between relative gene set expression in the brain and mean gsPRS importance scores were dependent on whether the gsPRS was calculated using the ADHD training subset or from summary statistics of the correlated phenotypes. In both groups (ADHD and correlated phenotypes), the correlation between gene set expression and importance scores remained significant but the correlation of relative gene set expression in the brain and gsPRS importance score was stronger when the gsPRS was developed using summary statistics from correlated phenotypes (see Figure 4). This finding may seem counterintuitive, considering that most of the gsPRS from other phenotypes had low gsPRS importance scores relative to the gsPRS calculated using the ADHD training subset.

However, when a gsPRS from a correlated phenotype is predictive in ADHD that gene set has shown an association and importance in its initial study, the ADHD training subset in our study, and the ADHD test subset in our study. We find it unsurprising that gsPRSs calculated from such generalizable gene sets would be more likely to represent true risk signals and therefore be more likely to have increased relative gene set expression values in the brain.

The learning curves suggest that the performance improvements from gsPRS should increase with increasing sample size. More complex models generally require more data to train, as demonstrated by the early stages of the learning curve that show perfect training subset performance and no predictability in the validation subset as the model is complex enough and sample size is low enough to memorize the training data instead of learning patterns among those data. In both learning curves, it is evident that the model is better at predicting the training data compared to the validation data even after selecting hyperparameters that specifically maximize prediction in the validation subset. This further illustrates the importance of testing performance on data the model does not learn from during training to get an accurate representation of model performance and generalization. As training size increases, the model can no longer rely on memorization and starts to learn patterns that generalize to the validation subset. The continued validation subset prediction improvements at the highest training sizes suggest that the model could still benefit from more training data. In comparison, the learning curve of a random forest model using only PRS-CS shows a quick plateau to optimal performance and additional training size does not further improve performance.

Our study has several limitations that could limit performance. To best estimate model performance and reduce overfitting, we split our data into several subsets, thereby limiting the number of study participants available to train the models. We also adjusted for the effects of the top 5 principal components in a PCA of the training subset to control for ancestry. This adjustment could inadvertently remove non-confounding information that might have improved performance and likely does not remove all ancestry information. A better method of selectively removing known confounders like ancestry would likely further improve both the performance and generalizability of these models. The gene sets we used to sum sets of SNPs into gsPRS, although capture the biological functions and pathways, may not be ideally suited for prediction tasks. A more data-driven approach to develop sets of SNPs that best collectively predict diagnosis may be necessary to maximize prediction performance.

More advanced machine learning methods and architectures may also lead to more predictive models. Including data beyond genotype information like clinical data and data that captures at least a portion of the environmental component of ADHD pathology could help machine learning models better estimate ADHD risk and better separate ADHD cases and controls. With the right set of interpretation tools, models that can accurately discriminate ADHD cases and controls would be useful in improving our understanding of the disorder and allow for testing specific hypotheses.

## Data Availability

Supplementary Data contains all information on data availability for summary statistics with links. Individual-level genotype data are available upon request to the Psychiatric Genomics Consortium (PGC).

https://atlas.ctglab.nl/ukb2_sumstats/f.2139.0.0_res.EUR.sumstats.MACfilt.txt.gz

http://cnsgenomics.com/data/wu_et_al_2019_nc/23_medication-taking_GWAS_summary_statistics.tar.gz

http://ssgac.org/documents/SSGAC_Rietveld2013.zip

http://ssgac.org/documents/CHIC_Summary_Benyamin2014.txt.gz

ftp://ftp.ebi.ac.uk/pub/databases/gwas/summary_statistics/Riveros-McKayF_30677029_GCST007241/SCOOP_UKHLS_ldcorrected.gz

https://www.med.unc.edu/pgc/results-and-downloads/downloads

https://atlas.ctglab.nl/ukb2_sumstats/f.1070.0.0_res.EUR.sumstats.MACfilt.txt.gz

https://www.med.unc.edu/pgc/results-and-downloads

http://enigma.ini.usc.edu/research/download-enigma-gwas-results/

https://atlas.ctglab.nl/ukb2_sumstats/f.3581.0.0_res.EUR.sumstats.MACfilt.txt.gz

## Acknowledgements

This project has received funding from the European Union’s Horizon 2020 research and innovation programme grant agreement No 667302. This project has received funding from the European Union’s Horizon 2020 research and innovation programme grant agreement No 965381.

The data used for the gene expression analyses described in this manuscript were obtained from the Genotype-Tissue Expression (GTEx) Portal. The GTEx Project was supported by the Common Fund of the Office of the Director of the National Institutes of Health, and by NCI, NHGRI, NHLBI, NIDA, NIMH, and NINDS.

## Supplementary Material

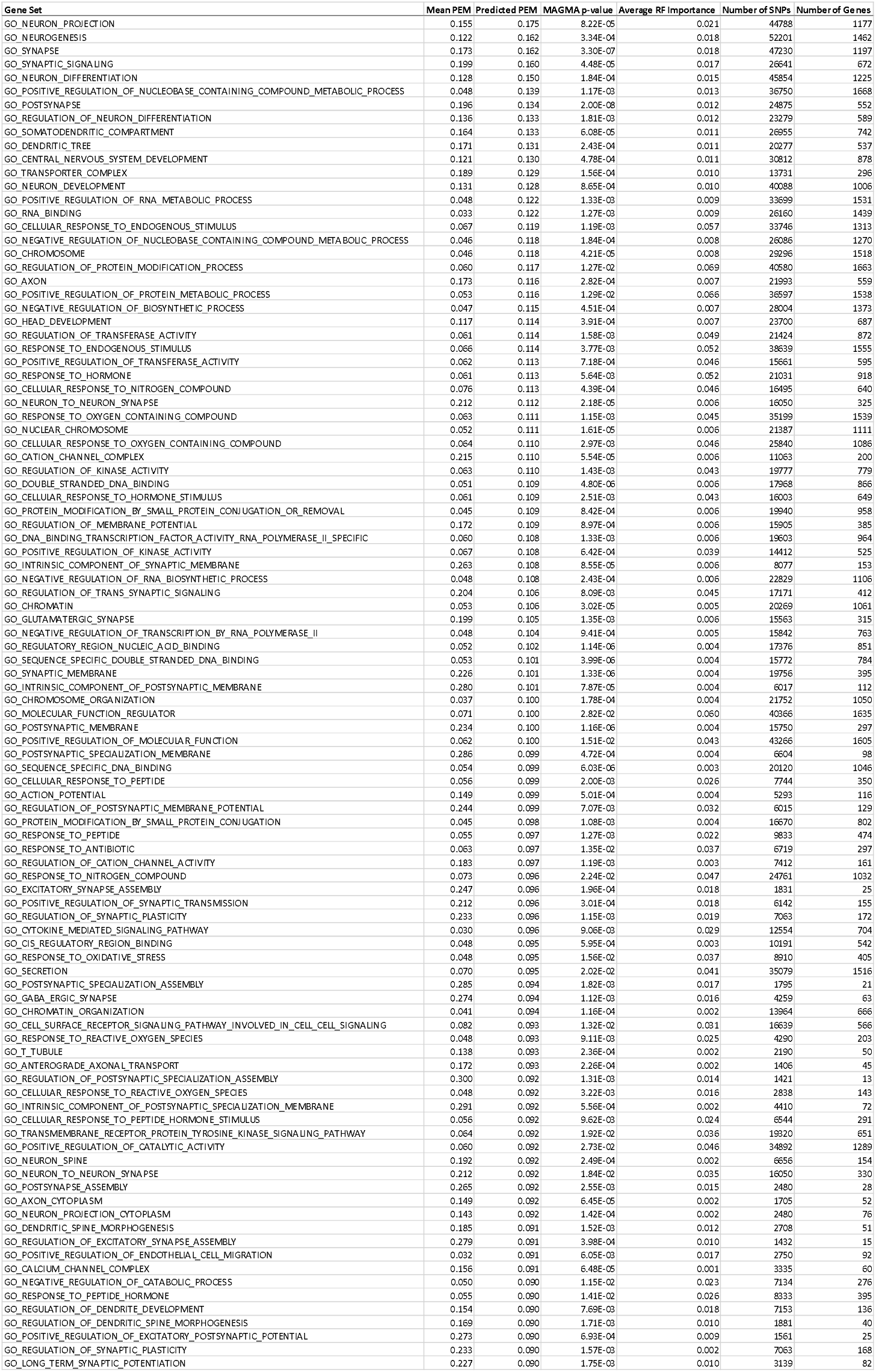

### Summary Statistics with Source Links

| ID | Accessibi<br>lity | Trait.x | rg | abs(rg) | se | z | P | P.bon | PMID | Year | uniqTrait | Popul<br>ation | Case | Control | N | SNP.h2 | File |
| --- | --- | --- | --- | --- | --- | --- | --- | --- | --- | --- | --- | --- | --- | --- | --- | --- | --- |
| 3304 | Free<br>download | Age first<br>had sexual<br>intercourse | -<br>0.584 | 0.584 | 0.031 | -18.938 | 0 | 0 | 31427789 | 2019 | Age first had<br>sexual<br>intercourse | UKB2<br>(EUR) | NA | NA | 339614 | 0.1132 | https://atlas.ctglab.nl/ukb2_sumstats/f.2139.0.0_res.EUR.sumstats.MACfilt.txt.gz |
| 4230 | Free<br>download | Opioids | 0.565 | 0.565 | 0.045 | 12.55 | 4.09E-36 | 5.05E-33 | 31015401 | 2019 | Opioids | UKB2<br>(EUR) | 22982 | 55826 | 78808 | 0.146 | http://cnsgenomics.com/data/wu_et_al_2019_nc/23_medication-taking_GWAS_summary_statistics.tar.gz |
| 58 | Free<br>download | College<br>completion | -<br>0.524 | 0.524 | 0.052 | -10.104 | 5.30E-24 | 6.54E-21 | 23722424 | 2013 | College<br>completion | EUR | NA | NA | 95427 | 0.105 | http://ssgac.org/documents/SSGAC_Rietveld2013.zip |
| 59 | Free<br>download | Childhood<br>IQ | -<br>0.461 | 0.461 | 0.089 | -5.163 | 2.42E-07 | 0.000299 | 23358156 | 2014 | Childhood IQ | EUR | NA | NA | 12441 | 0.2744 | http://ssgac.org/documents/CHIC_Summary_Benyamin2014.txt.gz |
| 4297 | Free<br>download | Extreme<br>obesity<br>(childhood) | 0.436 | 0.436 | 0.071 | 6.136 | 8.45E-10 | 1.04E-06 | 30677029 | 2019 | Extreme<br>obesity | EUR | 1456 | 6460 | 7916 | 0.5078 | ftp://ftp.ebi.ac.uk/pub/databases/gwas/summary_statistics/Riveros-McKayF_30677029_GCST007241/SCOOP_UKHLS_ldcorrected.gz |
| 4037 | Free<br>download | Autism<br>spectrum<br>disorder | 0.384 | 0.384 | 0.053 | 7.193 | 6.35E-13 | 7.84E-10 | 30804558 | 2019 | Autism<br>spectrum<br>disorder | EUR | 18381 | 27969 | 46350 | 0.1944 | https://www.med.unc.edu/pgc/results-and-downloads/downloads |
| 3219 | Free<br>download | Time spent<br>watching<br>television<br>(TV) | 0.372 | 0.372 | 0.032 | 11.766 | 5.86E-32 | 7.24E-29 | 31427789 | 2019 | Time spent<br>watching<br>television | UKB2<br>(EUR) | NA | NA | 365236 | 0.1023 | https://atlas.ctglab.nl/ukb2_sumstats/f.1070.0.0_res.EUR.sumstats.MACfilt.txt.gz |
| 1191 | Free<br>download | PGC cross<br>disorder | 0.262 | 0.262 | 0.053 | 4.954 | 7.28E-07 | 0.000898 | 23453885 | 2013 | PGC cross<br>disorder | EUR | 33332 | 27888 | 61220 | 0.1715 | https://www.med.unc.edu/pgc/results-and-downloads |
| 1226 | Free<br>download | Intracranial<br>Volume | -<br>0.248 | 0.248 | 0.058 | -4.314 | 1.60E-05 | 0.0198 | 27694991 | 2016 | Intracranial<br>Volume | EUR | NA | NA | 26577 | 0.2467 | http://enigma.ini.usc.edu/research/download-enigma-gwas-results/ |
| 3366 | Free<br>download | Age at<br>menopaus<br>e (last<br>menstrual<br>period)<br>(female) | -<br>0.226 | 0.226 | 0.039 | -5.844 | 5.08E-09 | 6.27E-06 | 31427789 | 2019 | Age at<br>menopause | UKB2<br>(EUR) | NA | NA | 119160 | 0.118 | https://atlas.ctglab.nl/ukb2_sumstats/f.3581.0.0_res.EUR.sumstats.MACfilt.txt.gz |
| 3539 | Free<br>download | Reason for<br>glasses/co<br>ntact<br>lenses: For<br>short-<br>sightednes<br>s, i.e. only<br>or mainly<br>for<br>distance<br>viewing<br>such as<br>driving,<br>cinema etc<br>(called<br>'myopia') | -<br>0.217 | 0.217 | 0.048 | -4.523 | 6.08E-06 | 0.0075 | 31427789 | 2019 | Reason for<br>glasses/contact<br>lenses: For<br>short-<br>sightedness | UKB2<br>(EUR) | 32082 | 46565 | 78647 | 0.1532 | https://atlas.ctglab.nl/ukb2_sumstats/6147_1_logistic.EUR.sumstats.MACfilt.txt.gz |

